# Immuno-fibrotic drivers of impaired lung function in post-acute sequelae of SARS-CoV-2 infection (PASC)

**DOI:** 10.1101/2021.01.31.21250870

**Authors:** Hyung J. Chun, Elias Coutavas, Alexander Pine, Alfred I. Lee, Vanessa Yu, Marcus Shallow, Coral X. Giovacchini, Anne Mathews, Brian Stephenson, Loretta G. Que, Patty J. Lee, Bryan D. Kraft

## Abstract

**Introduction:** Subjects recovering from COVID-19 frequently experience persistent respiratory ailments; however, little is known about the underlying biological factors that may direct lung recovery and the extent to which these are affected by COVID-19 severity.

**Methods:** We performed a prospective cohort study of subjects with persistent symptoms after acute COVID-19, collecting clinical data, pulmonary function tests, and plasma samples used for multiplex profiling of inflammatory, metabolic, angiogenic, and fibrotic factors.

**Results:** Sixty-one subjects were enrolled across two academic medical centers at a median of 9 weeks (interquartile range 6-10) after COVID-19 illness: n=13 subjects (21%) mild/non-hospitalized, n=30 (49%) hospitalized/non-critical, and n=18 subjects (30%) hospitalized/intensive care (“ICU”). Fifty-three subjects (85%) had lingering symptoms, most commonly dyspnea (69%) and cough (58%). Forced vital capacity (FVC), forced expiratory volume in 1 second (FEV1), and diffusing capacity for carbon monoxide (DLCO) declined as COVID-19 severity increased (P<0.05), but did not correlate with respiratory symptoms. Partial least-squares discriminant analysis of plasma biomarker profiles clustered subjects by past COVID-19 severity. Lipocalin 2 (LCN2), matrix metalloproteinase-7 (MMP-7), and hepatocyte growth factor (HGF) identified by the model were significantly higher in the ICU group (P<0.05) and inversely correlated with FVC and DLCO (P<0.05), and were confirmed in a separate validation cohort (n=53).

**Conclusions:** Subjective respiratory symptoms are common after acute COVID-19 illness but do not correlate with COVID-19 severity or pulmonary function. Host response profiles reflecting neutrophil activation (LCN2), fibrosis signaling (MMP-7), and alveolar repair (HGF) track with lung impairment and may be novel therapeutic or prognostic targets.

**Funding:** The study was funded in part by the NHLBI (K08HL130557 to BDK and R01HL142818 to HJC), the DeLuca Foundation Award (AP), a donation from Jack Levin to the Benign Hematology Program at Yale, and Divisional/Departmental funds from Duke University.

## INTRODUCTION

As the number of patients who recover from acute coronavirus disease 19 (COVID-19) rises, it is increasingly apparent that a substantial subset displays persistent subjective and objective respiratory ailments. Patients report a high prevalence of lingering respiratory symptoms such as fatigue and dyspnea (1-4), and spirometry and diffusion capacity testing show impaired pulmonary function (3). However, little is known about the biological drivers of long-term respiratory disease after COVID-19, and how acute COVID illness severity impacts the emerging COVID-19 long-hauler syndrome, or post-acute sequelae of SARS-CoV-2 infection (PASC). To better understand the relationship between subjective and objective respiratory abnormalities and underlying biological drivers, we measured symptom burden, pulmonary function tests, and plasma biomarkers in subjects that have recovered from COVID-19 infection.

## RESULTS

We enrolled 61 subjects into the post-COVID-19 clinics. Subject demographics for each group are shown in the **Table**. Median (IQR) age was 53 (43-62) years and sex distribution was equitable: 56% males and 44% females. Median (IQR) time from onset of COVID-19 infection to post-COVID follow up was 9 (6-11) weeks, but was not significantly different between the three COVID-19 groups. Thirteen subjects (21%) recovered at home (“home” group), 30 subjects (48%) were hospitalized but did not require ICU (“non-ICU” group) and nineteen subjects (31%) were hospitalized and required ICU level care (“ICU” group). Eight of the subjects admitted to the ICU required intubation and mechanical ventilation, but none required tracheostomy. The demographics and characteristics of the subgroup of subjects that underwent plasma biomarker testing are also shown.

Fifty-three subjects (85%) reported lingering symptoms of COVID-19, the most common being dyspnea (69%), cough (58%), fever (47%), fatigue (29%), chest pain (26%), and headache (23%) (**Figure 1A**). There were no significant differences in symptom burden between the home, non-ICU, and ICU groups (P=NS, **Figure 1B**). Pulmonary function testing was performed on 59 subjects. Forced vital capacity (FVC), forced expiratory volume in 1 second (FEV1), and diffusing capacity for carbon monoxide (DLCO) decreased as severity of COVID-19 increased (**Figure 2A**). There was no relationship between pulmonary function and presence of persistent respiratory symptoms, measured by the sum of the scores for fatigue, dyspnea, and cough (**Figure 2B**).

**Figure 1:**
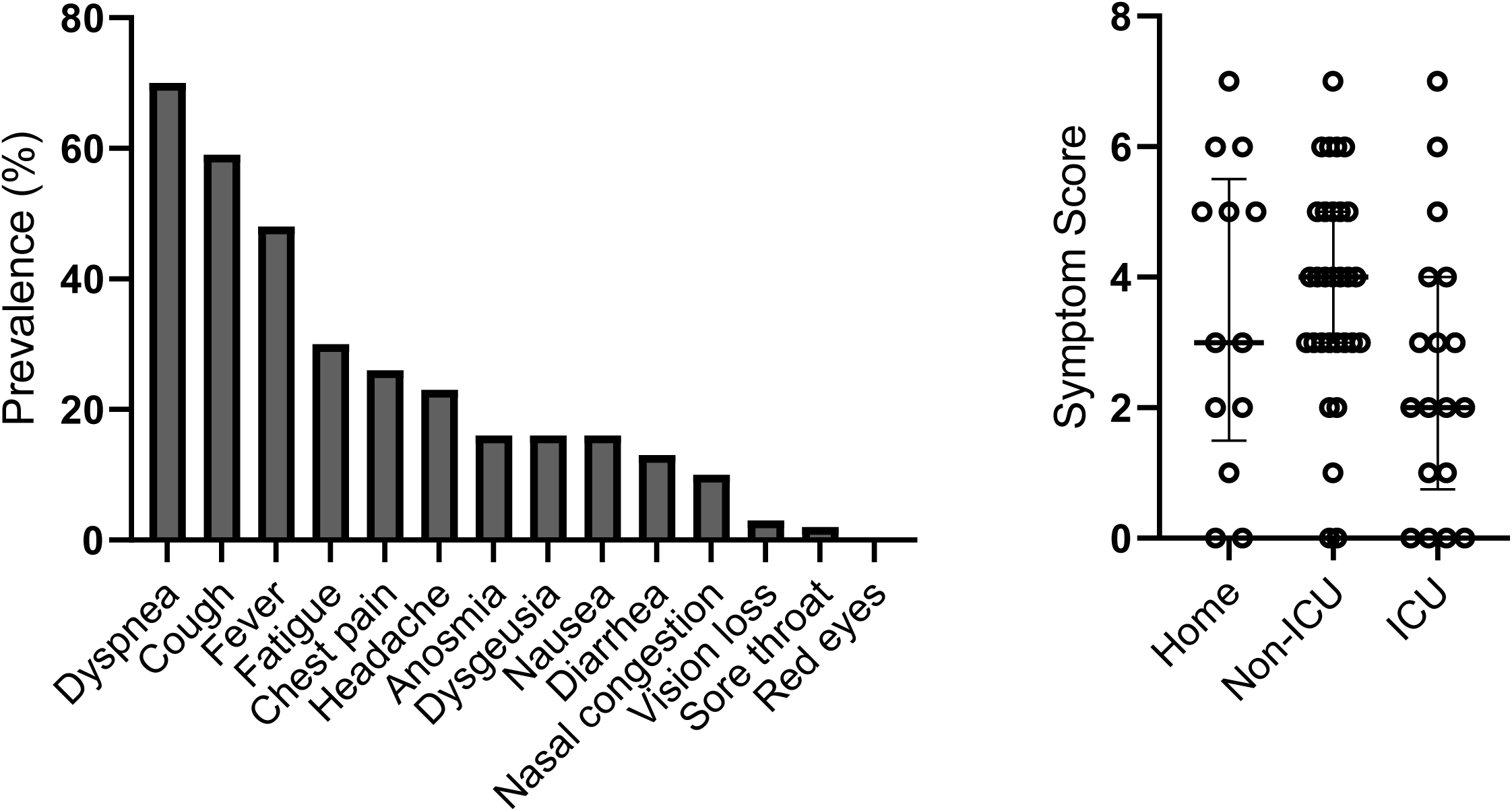
Post-COVID-19 symptom burden. A) Prevalence of symptoms reported by subjects (n=61) at the time of initial follow up visit. B) Symptom scores (total number of symptoms reported) with median bars and interquartile range error bars for home (n=13), non-ICU (n=30), and ICU (n=18) groups. There were no significant differences between groups using the Kruskal-Wallis test with Benjamini-Hochberg post-hoc test.

**Figure 2:**
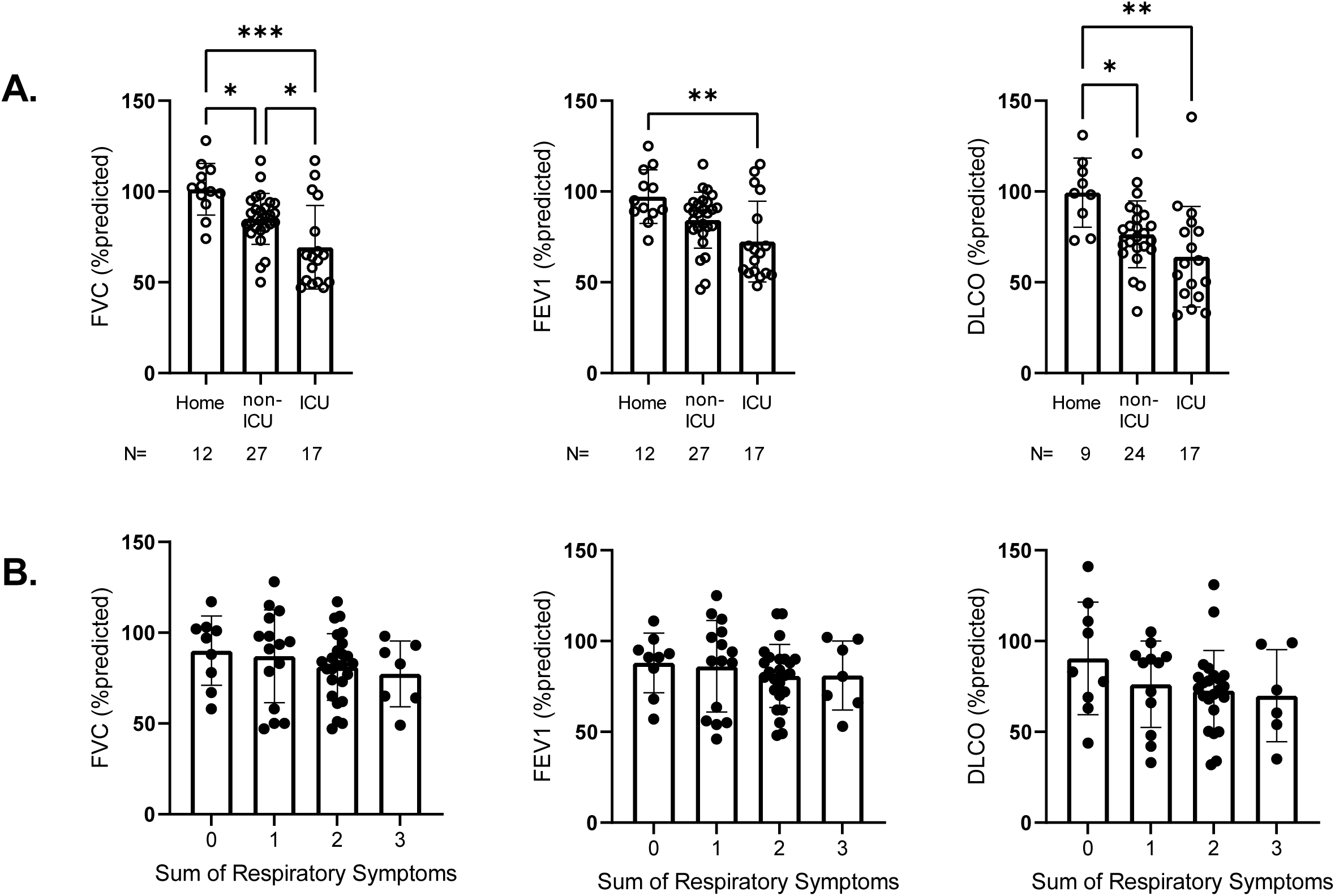
Pulmonary function tests and Respiratory Symptom Score. A) Scatter plots show forced vital capacity (FVC), forced expiratory volume in 1 second (FEV1), and diffusing capacity for carbon monoxide (DLCO) as percentages of predicted values for each subject in the home, non-ICU, and ICU groups for N (number) shown. B) Scatter plots show the sum of respiratory symptoms (1 point each for dyspnea, cough, and fatigue) vs. FVC, FEV1, and DLCO. Bars are medians and error bars are interquartile range. Statistical analysis by Kruskal-Wallis test with Benjamini-Hochberg post-hoc test. *P<0.05, **P<0.01, ***P<0.001.

To identify potential biological factors that associate with impaired pulmonary function after COVID-19 infection, we measured levels of circulating inflammatory, metabolic, angiogenic, and fibrotic markers in plasma from a subset of 22 subjects (home group n=7, non-ICU group n=5, ICU group n=10) and performed partial least-squares discriminant analysis (PLS-DA) (**Figure 3**). The score plot (**Figure 3A**) shows that subjects largely cluster in distinct quadrants based on how severe their COVID-19 illness was, even ∼9 weeks after infection. In fact, the clustering shows a counter-clockwise rotation around the origin as severity of viral infection increases from home to non-ICU to ICU-level of care. The loadings plot (**Figure 3B**) shows the associations between the clusters of subjects and the measured plasma biomarkers (**Supplemental Table**). The top right quadrant shows the biomarkers that directly correlate with critical COVID-19 illness requiring ICU admission. Several of the most impactful biomarkers (i.e. furthest from the point of origin) that associate with ICU level of care are LCN2 (lipocalin-2), MMP-7 (matrix metalloproteinase-7), and HGF (hepatocyte growth factor). Plasma levels of all three factors were similar between the control and home groups, but increased with COVID-19 severity, and were significantly higher in the ICU group (**Figure 4A**).

**Figure 3:**
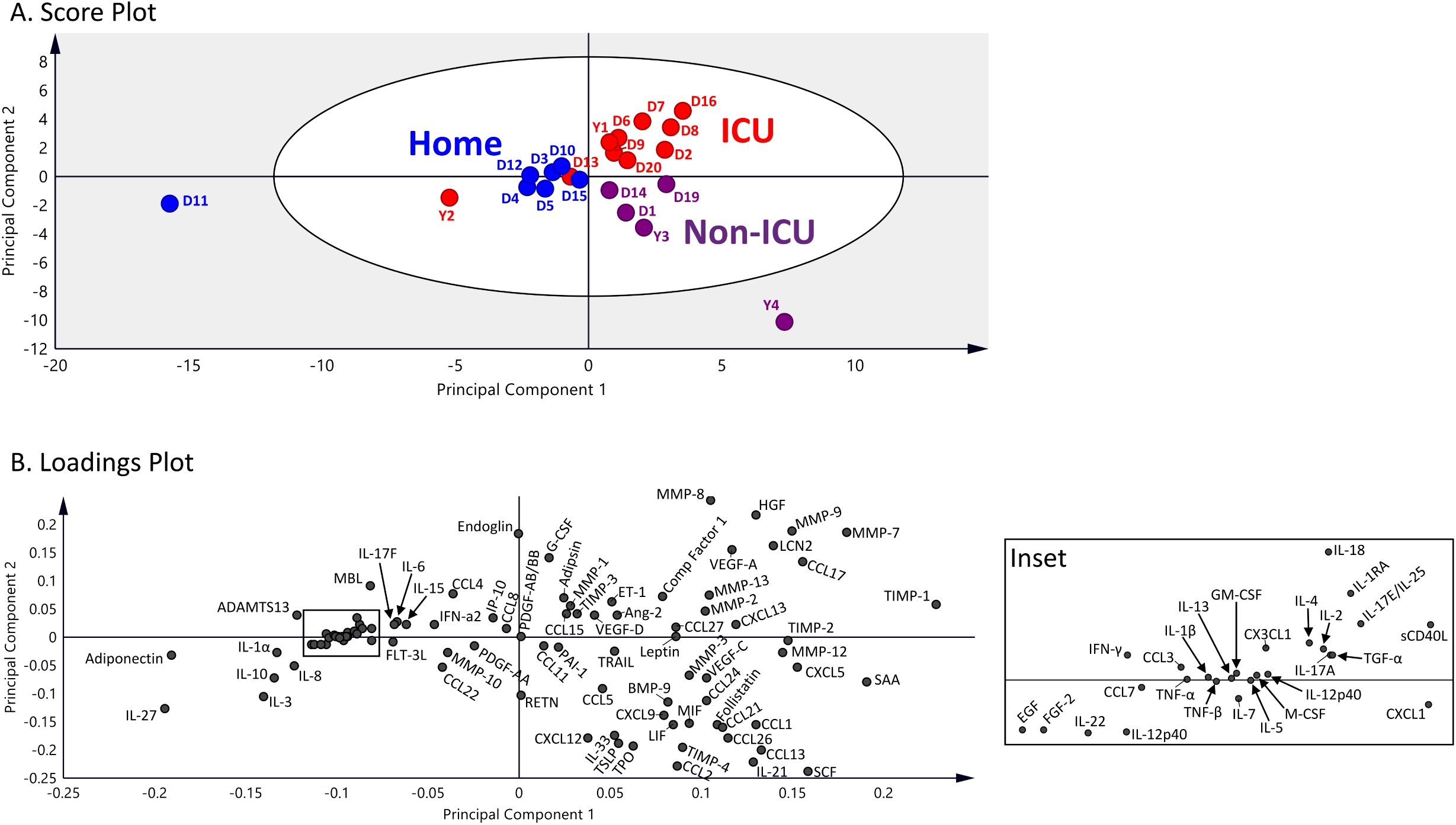
Partial least-squares discriminant analysis (PLS-DA). A) The score plot shows each individual subject in the home (blue circles), non-ICU (purple circles), and ICU (red circles) groups. The grey zone in the score plot depicts outliers with 95% confidence. B) The loadings plot depicts associations between clusters and the measured biomarkers.

**Figure 4:**
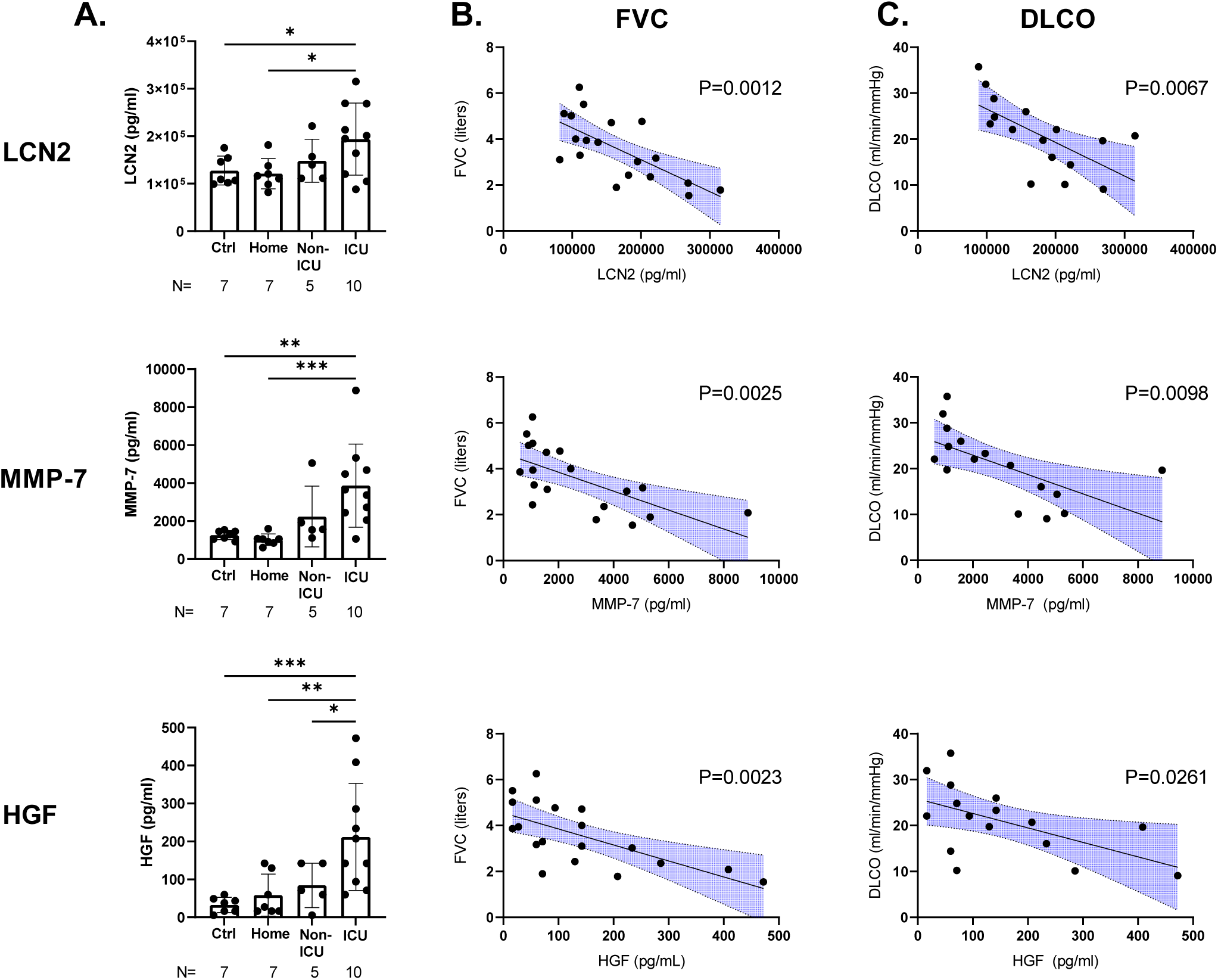
Biomarkers. A) Plasma levels of LCN2 (lipocalin-2), MMP-7 (matrix metalloproteinase-7), and HGF (hepatocyte growth factor) in individual subjects (black circles) in the control (Ctrl), home, non-ICU, and ICU groups. Bars are means and error bars are standard deviation. Statistical analysis by one-way ANOVA with Fisher’s LSD post-hoc test. *P<0.05, **P<0.01, ***P<0.001. B). Scatter plots for subjects (black circles) of LCN2 (lipocalin-2), MMP-7 (matrix metalloproteinase-7), and HGF (hepatocyte growth factor) on abscissa versus FVC (forced vital capacity) or DLCO (diffusing capacity) on ordinate. Statistical analysis is simple linear regression. Blue shaded area shows 95% confidence intervals.

To further determine if specific circulating factors are associated with persistently impaired pulmonary function, we used random-forest feature selection (5) to identify which factors best explain the variability seen in FVC and DLCO (**Supplemental Figures 1-2**). This analysis identified MMP-7 as the marker that was most significantly associated with FVC and DLCO. LCN2, HGF, and leptin, among others, were also significantly associated with FVC, DLCO, or both. In our patient cohort, leptin did not differ by acute COVID-19 severity, but was strongly inversely correlated with post-COVID-19 FVC and also directly correlated with body mass index (**Supplemental Figure 3**). However, after adjusting for body mass index, the relationship between leptin and FVC was no longer significant. Confirmation of the relationship between LCN2, MMP-7, and HGF and pulmonary function using simple linear regression identified a highly significant, inverse correlation with FVC and DLCO (**Figure 4B and 4C**). There was no relationship between these three biomarkers and burden of subjective symptoms.

To validate these three biomarkers, we measured them in a separate group of 53 subjects (“validation cohort”) (**Table** and **Supplemental Figure 4A**). The validation cohort consisted of n=13 subjects in the home group, n=31 subjects in the hospitalized, non-ICU group, and n=9 subjects in the ICU group. Demographics of the discovery and validation cohorts were comparable (**Table**). Plasma levels of LCN2 and MMP-7 were significantly higher in the validation ICU group compared with the validation home group, whereas a statistical trend was observed for HGF (**Supplemental Figure 4A**). However, like the discovery cohort, all three biomarkers were highly linearly correlated with decline in FVC and DLCO (**Supplemental Figures 4B and 4C**).

## DISCUSSION

Herein we report that among patients returning for follow-up visits for persistent symptoms after their acute COVID-19 illness, respiratory symptoms and impaired lung function are common after acute COVID-19 infection. Pulmonary function in particular worsens as COVID-19 illness severity increases. We further show that plasma biomarker profiles measured at 9 weeks after COVID-19 infection reveal prior COVID-19 severity and cluster patients accurately. Plasma LCN-2, MMP-7, and HGF levels are highest in survivors of severe COVID-19 infection and correlate strongly with pulmonary function impairment.

Patients recovering from acute COVID-19 infection commonly display persistent symptoms such as fatigue and dyspnea (1-4). Our study found a high degree of symptom burden across subgroups (home, non-ICU, ICU) that did not associate by COVID-19 severity (3). Impaired pulmonary function was reported by Huang et al (1), and similar to our findings, correlated with prior COVID-19 severity. Together these data highlight the discordance of subjective and objective respiratory abnormalities after COVID-19 infection (3), a well-known phenomenon in cardiopulmonary diseases in general (6), and indicate that subjective symptoms alone cannot discern degree of post-COVID-19 lung function impairment.

Partial least-squares discriminant analysis could reliably identify and separate subjects by COVID-19 illness severity based on their plasma biomarker profiles, even 9 weeks after recovery. Our analysis highlighted a number of important biomarkers that strongly associated with severe/ICU COVID-19 status. We chose to further analyze three of these markers, LCN2, MMP-7, and HGF based on their differing roles in host response. While these markers were similar between the healthy controls and the home group, there was a stepwise increase seen as COVID-19 severity increased. Furthermore, LCN2, MMP-7, and HGF were not associated with subjective symptoms in this study, or reported elsewhere to associate with subjective symptoms, but were strongly associated with objective respiratory function. These findings are confirmed by the validation cohort, which also found strong linear relationships between plasma biomarker levels and decline in respiratory function. LCN2 is a component of neutrophil secondary granules that was found to strongly associate with acute COVID-19 critical illness and mortality (7-10); however, the role in post-COVID-19 recovery has not been previously described. That higher levels of LCN-2 are found at 9 weeks after severe COVID-19 illness compared with non-severe cases suggests these patients may have ongoing neutrophil activation that could be amenable to targeted therapy.

MMP-7 is a protease that breaks down extracellular matrix deposited in the lung after injury. The role of MMP-7 in COVID-19 has not been reported; however, previous studies of non-COVID-19 acute lung injury have shown higher circulating levels of MMPs (11) that associate with worse disease severity and clinical outcomes (12). MMP-7 is also a validated biomarker for idiopathic pulmonary fibrosis severity that correlates with pulmonary function (13), and our findings suggest MMP-7 may be a novel biomarker of COVID-19 ARDS recovery.

HGF is a growth factor secreted by alveolar fibroblasts shown to be elevated in bronchoalveolar lavage fluid of patients with ARDS (14) and in plasma of patients with acute COVID-19 illness (15); however, the plasma levels after COVID-19 recovery have not been previously reported. In vivo and in vitro studies indicate HGF promotes alveolar epithelial and endothelial repair after acute lung injury (16, 17) via induction of counter inflammatory and anti-oxidant gene expression (18-20). Our findings suggest high HGF levels at 9 weeks post-COVID-19 could indicate ongoing alveolar repair proportionate to the degree of acute lung injury; however, further studies are needed to determine the impact of HGF on COVID-19 recovery.

Leptin is a hormone secreted by adipocytes in proportion to obesity, a risk factor for severe acute COVID-19 (21), and may contribute to immune dysregulation during acute COVID-19 (22, 23). In our patient population with post-COVID-19 syndrome, we found leptin was inversely related to FVC, however, after adjusting for body mass index, the relationship was no longer significant, indicating body mass index was a confounder. Future studies should take body mass index into account when reporting leptin results.

Our study has several limitations. First, we only recruited subjects that had ongoing symptoms after COVID-19 infection, therefore we cannot report the true prevalence of symptoms or pulmonary function derangements in subjects with asymptomatic infection or in subjects with symptom resolution after COVID-19 infection. Second, our plasma biomarker profiles represent a single time point for each subject, and therefore we cannot draw conclusions about trajectory or changes over time. Third, we cannot establish mechanism in this human cohort study; however, our findings are highly associational and lay the foundations to drive further work in this area. Fourth, the PLS-DA model was at risk for being over-fit, however this was mitigated with similar findings of the Boruta algorithm, and addressed with the addition of the validation cohort which confirmed the findings of the discovery cohort. Lastly, the majority of the patients were assessed at 9 weeks after their acute COVID-19 infection, which may not provide sufficient insights into the long term lung repair that is likely to occur beyond this timeframe. Future studies evaluating biomarker profiles and pulmonary function at later time points will provide greater insights into persistent symptoms associated with COVID-19 illness.

In conclusion, persistent respiratory symptoms and impaired lung function are common after acute COVID-19 illness, but subjective symptoms alone do not predict lung injury. Partial least-squares discriminant analysis reliably identifies and separates subjects by COVID-19 illness severity based on their plasma biomarker profiles, even 9 weeks after recovery. Circulating factors associated with acute neutrophil activation, fibrosis signaling, and alveolar epithelial repair remain elevated in survivors of acute COVID-19 infection and strongly associate with impaired pulmonary function. Our study provides novel insights into divergent host responses of lung repair after COVID-19 pneumonia and suggest that immunologic and fibrotic drivers of disease severity in the acute COVID-19 illness may regulate the trajectory of recovery in the post-COVID-19 phase. These markers may represent key prognostic tools that can be followed during and after COVID-19 illness, and may identify COVID-19 survivors at increased risk for chronic parenchymal lung disease, and will require validation in a larger study.

## METHODS

### Subject Enrollment

The studies were approved by the institutional review boards of Yale University (#2000027792) and Duke University (Pro00105518 and Pro00106151). Subjects with persistent symptoms 30 days after the diagnosis of acute COVID-19 infection were considered eligible and recruited to a post-COVID-19 clinic for further evaluation. Seven asymptomatic, non-hospitalized healthy control subjects were enrolled at Yale under a separate approved protocol (#1401013259) (median age 45 (interquartile range 44-53), 2 male, 5 female). Written informed consent was provided by all subjects prior to study activities.

### Data Collection

All subjects completed symptom questionnaires (presence or absence of the following: fatigue, fever, headache, dyspnea, cough, chest pain, nausea, diarrhea, anosmia, dysgeusia, sore throat, nasal congestion, red eyes, and vision loss), physical examinations, and pulmonary function testing (spirometry and diffusion capacity) at the time of their follow-up visits. Symptom scores were calculated by the sum of the number of symptoms reported. Respiratory symptom scores were calculated by the sum of the following symptoms: cough, shortness of breath, and fatigue. Severity of COVID-19 infection was defined by need for intensive care unit (ICU) hospital care, non-ICU hospital care (non-ICU), or non-hospitalized home recuperation (home). Whole blood was collected by venipuncture, centrifuged, separated into plasma, and stored at - 80°C until use.

### Plasma biomarker profiling

Multiplex profiling of plasma was performed on a subset of discovery subjects (n=22) and a validation cohort (n=53) by Eve Technologies (Calgary, Alberta, Canada) using the following assays: Human Cytokine 71-Plex, Human Complement Panel 1, Human Angiogenesis 17-Plex, Human MMP 9-Plex and TIMP 4-Plex, and Human Adipokine 5-Plex. A complete list of biomarkers profiled is provided in the **Supplemental Table**.

### Statistical Analysis

Grouped data are expressed as median (interquartile range) unless specified otherwise. Differences between groups were analyzed by Kruskal-Wallis test with Benjamini-Hochberg post-hoc test to control for false discovery rate or one-way ANOVA with Fisher’s LSD post-hoc test (Graphpad Prism, San Diego, CA). Simple linear regression and multivariate linear regression were performed by Prism. Skewed datasets were log-transformed for normality. Partial least-squares discriminant analysis (PLS-DA) was performed using SIMCA v15 (Umetrics, Umeå, Sweden). The random-forest feature selection (Boruta algorithm) was performed using the Boruta package in R software. All P-values are two-tailed. P<0.05 was accepted as statistically significant. Statistical trends were noted for P<0.1.

## Data Availability

All data referred to in the manuscript is available.

## AUTHOR CONTRIBUTIONS

H.J.C. and B.D.K. wrote the manuscript that was approved by all authors. All authors participated in data collection, analysis, and/or interpretation. The overall study design was conceived by Y.J.C., B.D.K., L.G.Q., A.I.L., and P.J.L.

## ACKNOWLEDGEMENTS

The authors thank Dr. Amanda Brucker and Dr. Shein-Chung Chow for their statistical expertise and assistance with analytical interpretation and manuscript preparation, and Antoinette Santoro for assistance with patient enrollment.

---

**Table:** Subject Demographics.

**Table: Subject Demographics**
|  | **Post-COVID-19 Clinic**<br>**Discovery Cohort**<br>**N=61** | **Discovery**<br>**Biomarker**<br>**Subgroup**<br>**N=22** | **Validation**<br>**Cohort**<br>**N= 53** |
| --- | --- | --- | --- |
| **Age (years), median (IQR)** | 53 (43-65) | 51 (41-62) | 53 (45-65) |
| **Sex, N (%)** |  |  |  |
| *Female* | 27 (44%) | 11 (50%) | 24 (45%) |
| *Male* | 34 (56%) | 11 (50%) | 29 (55%) |
| **Ethnicity, N (%)** |  |  |  |
| *Hispanic/Latino* | 10 (16%) | 1 (5%) | 42 (79%) |
| *Not Hispanic/Latino* | 51 (84%) | 21 (95%) | 11 (21%) |
| **Race, N (%)** |  |  |  |
| *African American* | 14 (23%) | 7 (32%) | 15 (28%) |
| *American Indian or Alaskan Native* | 2 (3%) | 1 (4%) | 1 (2%) |
| *Asian* | 3 (5%) | 1 (4%) | 1 (2%) |
| *Caucasian* | 41 (67%) | 12 (56%) | 31 (58%) |
| *Two or more races* | 1 (2%) | 1 (4%) | 1 (2%) |
| *Other* | 0 (0%) | 0 (0%) | 4 (8%) |
| **Body mass index, median (IQR)** | 30 (26-36) | 27 (24-33) | 31 (29-39) |
| **COVID-19 Severity, N (%)** |  |  |  |
| *Home* | 13 (21%) | 7 (32%) | 13 (25%) |
| *Hospitalized, non-ICU* | 30 (49%) | 5 (23%) | 31 (58%) |
| *Hospitalized, ICU* | 18 (30%) | 10 (45%) | 9 (17%) |
| **Mechanical Ventilation, N (%)** | 8 (13%) | 4 (18%) | 7 (13%) |
| **Venovenous ECMO, N (%)** | 0 (0%) | 0 (0%) | 3 (6%) |
| **Weeks since COVID-19, median (IQR)** | 9 (6-10) | 6 (6-12) | 6 (6-6) |

*Abbreviations:* COVID-19, coronavirus disease 2019; ECMO, extracorporeal membrane oxygenation; ICU, intensive care unit; IQR, interquartile range; N, number.

## REFERENCES

1. Huang C, Huang L, Wang Y, Li X, Ren L, Gu X, et al. 6-month consequences of COVID-19 in patients discharged from hospital: a cohort study. Lancet. 2021;397(10270):220–32.

2. Carfi A, Bernabei R, Landi F, and Gemelli Against C-P-ACSG. Persistent Symptoms in Patients After Acute COVID-19. JAMA : the journal of the American Medical Association. 2020.

3. Townsend L, Dyer AH, Jones K, Dunne J, Mooney A, Gaffney F, et al. Persistent fatigue following SARS-CoV-2 infection is common and independent of severity of initial infection. PloS one. 2020;15(11):e0240784.

4. Alwan NA, Attree E, Blair JM, Bogaert D, Bowen MA, Boyle J, et al. From doctors as patients: a manifesto for tackling persisting symptoms of covid-19. Bmj. 2020;370:m3565.

5. Kursa MB, and Rudnicki WR. Feature selection with the Boruta package. J Stat Softw. 2010;36(11):1–13.

6. Leiner GC, Abramowitz S, Lewis WA, and Small MJ. Dyspnea and pulmonary function tests. The American review of respiratory disease. 1965;92(5):822–3.

7. Meizlish ML, Pine AB, Bishai JD, Goshua G, Nadelmann ER, Simonov M, et al. A neutrophil activation signature predicts critical illness and mortality in COVID-19. medRxiv. 2020:2020.09.01.20183897.

8. Overmyer KA, Shishkova E, Miller IJ, Balnis J, Bernstein MN, Peters-Clarke TM, et al. Large-Scale Multi-omic Analysis of COVID-19 Severity. Cell Syst. 2020.

9. Li G, Wang J, He X, Zhang L, Ran Q, Xiong A, et al. An integrative analysis identifying transcriptional features and key genes involved in COVID-19. Epigenomics. 2020;12(22):1969–81.

10. Abers MS, Delmonte OM, Ricotta EE, Fintzi J, Fink DL, de Jesus AAA, et al. An immune-based biomarker signature is associated with mortality in COVID-19 patients. JCI Insight. 2021;6(1).

11. O’Kane CM, McKeown SW, Perkins GD, Bassford CR, Gao F, Thickett DR, et al. Salbutamol up-regulates matrix metalloproteinase-9 in the alveolar space in the acute respiratory distress syndrome. Critical care medicine. 2009;37(7):2242–9.

12. Davey A, McAuley DF, and O’Kane CM. Matrix metalloproteinases in acute lung injury: mediators of injury and drivers of repair. The European respiratory journal. 2011;38(4):959–70.

13. Rosas IO, Richards TJ, Konishi K, Zhang Y, Gibson K, Lokshin AE, et al. MMP1 and MMP7 as potential peripheral blood biomarkers in idiopathic pulmonary fibrosis. PLoS Med. 2008;5(4):e93.

14. Quesnel C, Marchand-Adam S, Fabre A, Marchal-Somme J, Philip I, Lasocki S, et al. Regulation of hepatocyte growth factor secretion by fibroblasts in patients with acute lung injury. American journal of physiology Lung cellular and molecular physiology. 2008;294(2):L334–43.

15. Yang Y, Shen C, Li J, Yuan J, Wei J, Huang F, et al. Plasma IP-10 and MCP-3 levels are highly associated with disease severity and predict the progression of COVID-19. The Journal of allergy and clinical immunology. 2020;146(1):119–27 e4.

16. Ito Y, Correll K, Schiel JA, Finigan JH, Prekeris R, and Mason RJ. Lung fibroblasts accelerate wound closure in human alveolar epithelial cells through hepatocyte growth factor/c-Met signaling. American journal of physiology Lung cellular and molecular physiology. 2014;307(1):L94–105.

17. Wang H, Zheng R, Chen Q, Shao J, Yu J, and Hu S. Mesenchymal stem cells microvesicles stabilize endothelial barrier function partly mediated by hepatocyte growth factor (HGF). Stem Cell Res Ther. 2017;8(1):211.

18. Okada M, Sugita K, Inukai T, Goi K, Kagami K, Kawasaki K, et al. Hepatocyte growth factor protects small airway epithelial cells from apoptosis induced by tumor necrosis factor-alpha or oxidative stress. Pediatr Res. 2004;56(3):336–44.

19. Garnier M, Gibelin A, Mailleux AA, Lecon V, Hurtado-Nedelec M, Laschet J, et al. Macrophage Polarization Favors Epithelial Repair During Acute Respiratory Distress Syndrome. Critical care medicine. 2018;46(7):e692–e701.

20. Kamimoto M, Mizuno S, Matsumoto K, and Nakamura T. Hepatocyte growth factor prevents multiple organ injuries in endotoxemic mice through a heme oxygenase-1-dependent mechanism. Biochemical and biophysical research communications. 2009;380(2):333–7.

21. Sacco V, Rauch B, Gar C, Haschka S, Potzel AL, Kern-Matschilles S, et al. Overweight/obesity as the potentially most important lifestyle factor associated with signs of pneumonia in COVID-19. PloS one. 2020;15(11):e0237799.

22. Wang J, Xu Y, Zhang X, Wang S, Peng Z, Guo J, et al. Leptin correlates with monocytes activation and severe condition in COVID-19 patients. J Leukoc Biol. 2021.

23. AbdelMassih AF, Fouda R, Kamel A, Mishriky F, Ismail HA, El Qadi L, et al. Single cell sequencing unraveling genetic basis of severe COVID19 in obesity. Obes Med. 2020;20:100303.

