## Supplemental Figures 1- 4 for "Immuno-fibrotic drivers of impaired lung function in post-acute sequelae of SARS-CoV-2 infection (PASC)"

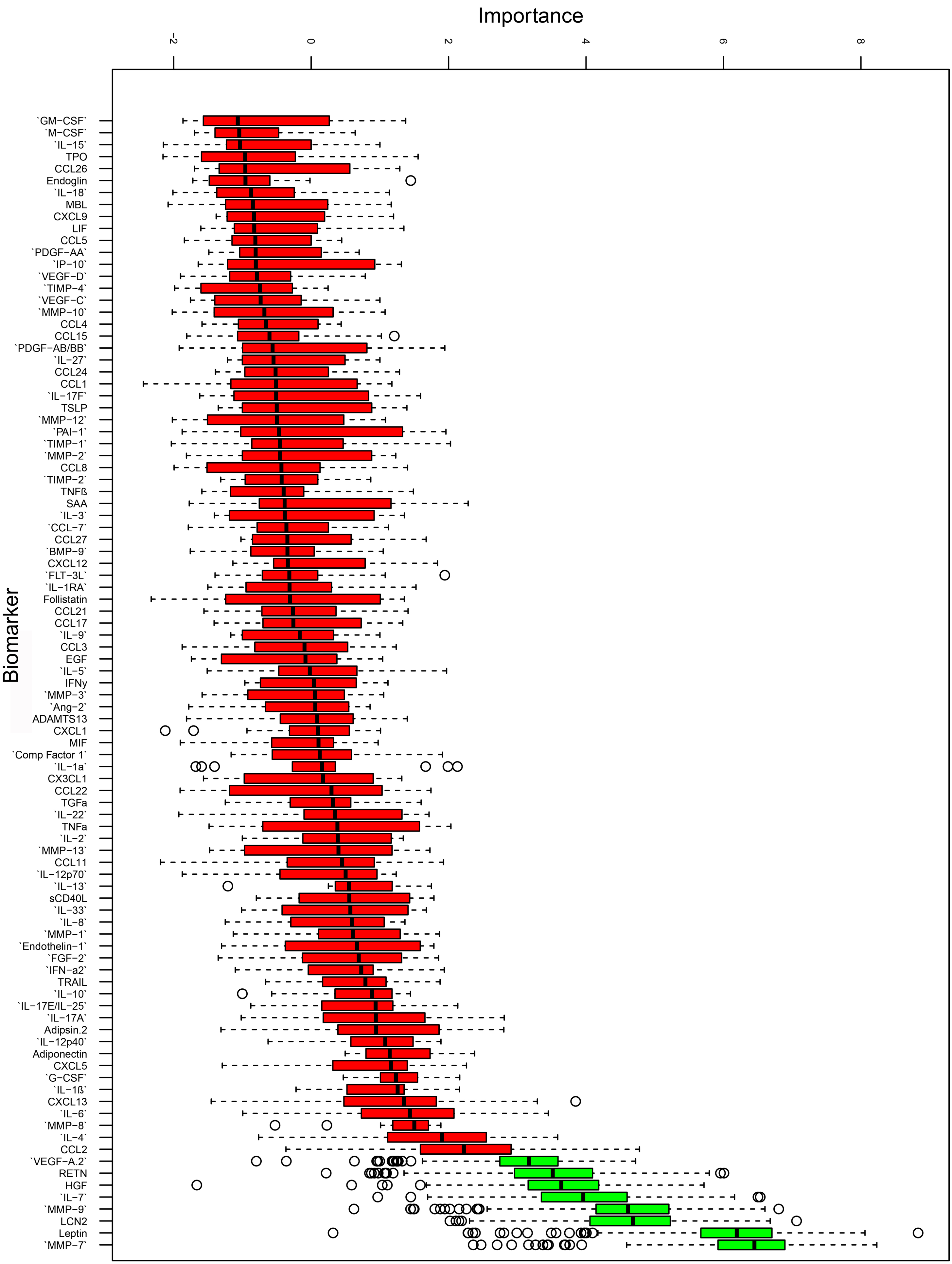


**Supplemental Figure 1:** Random-forest feature selection Boruta procedure showing the biomarkers in reverse order of importance for explaining variation in forced vital capacity. Green biomarkers are confirmed and red biomarkers are rejected.

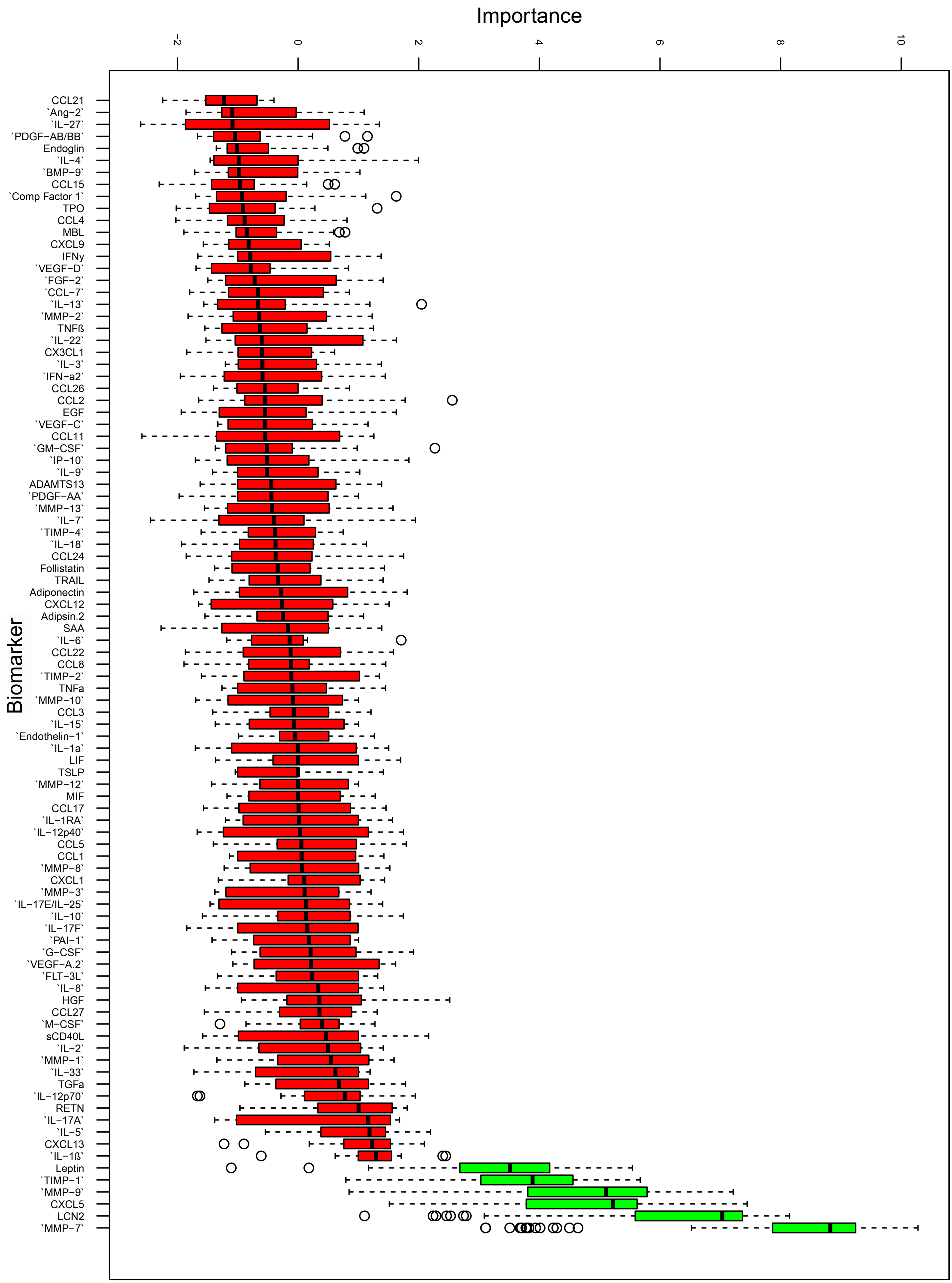


**Supplemental Figure 2:** Random-forest feature selection Boruta procedure showing the biomarkers in reverse order of importance for explaining variation in diffusion capacity for carbon monoxide. Green biomarkers are confirmed and red biomarkers are rejected.

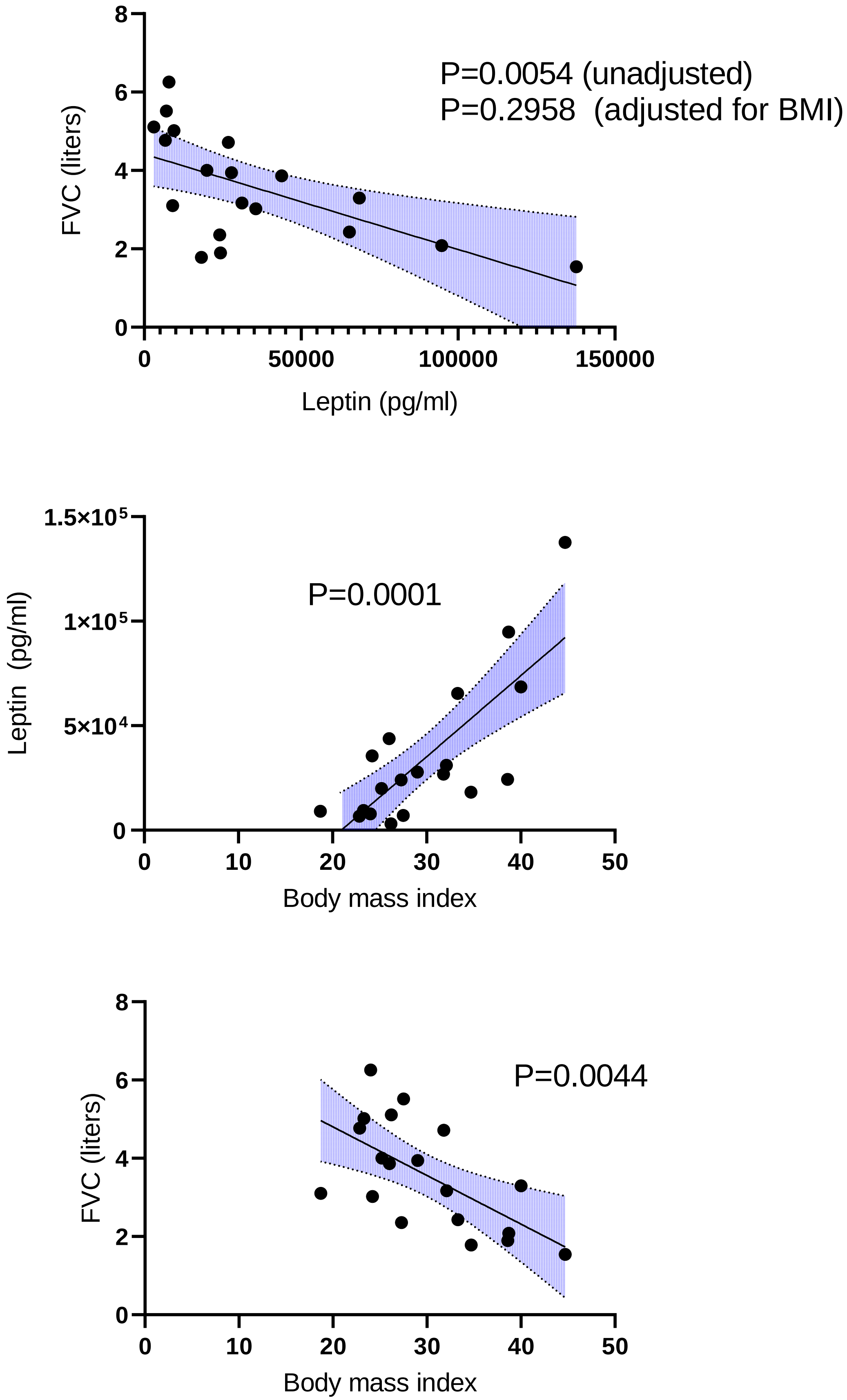


**Supplemental Figure 3: Leptin.** *Top*: Scatter plot of plasma leptin levels on abscissa and forced vital capacity (FVC) on ordinate for n=19 subjects. *Middle*: Scatter plot of body mass index (BMI) on abscissa and plasma leptin levels on ordinate for same 19 subjects. *Bottom*: Scatter plot of body mass index on abscissa and FVC on ordinate for same 19 subjects. Statistical analysis is multiple variable linear regression (Top) and simple linear regression (Middle and Bottom). Blue shaded regions show 95% confidence intervals.

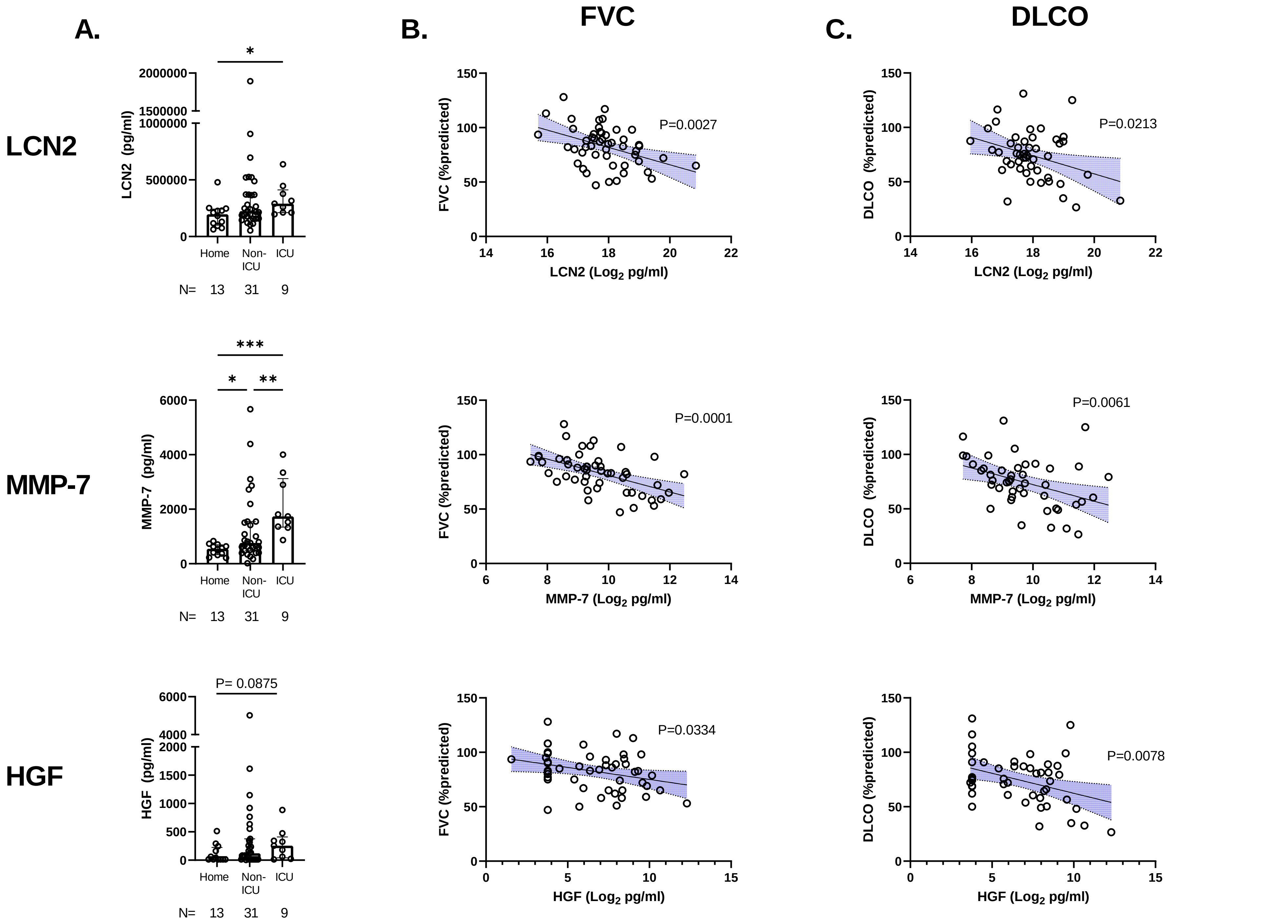


**Supplemental Figure 4: Validation Cohort biomarkers.** A) Plasma levels of lipcalin-2 (LCN2), MMP-7 (matrix metalloproteinase-7), and HGF (hepatocyte growth factor) in individual subjects (open circles) in the home, non-ICU, and ICU groups. Bars are medians with interquartile ranges. Statistical analysis by Kruskal-Wallis test with Dunn's post-hoc test. *P<0.05, **P<0.01, and ***P<0.001. B) Scatter plots for subjects (open circles) of LCN2, MMP-7, and HGFon abscissa versus percent predicted FVC (forced vital capacity) and DLCO (diffusing capacity) on ordinate. Biomarker levels are Log2 transformed for normality. Statistical analysis is simple linear regression. Blue shaded area shows 95% confidence intervals.
